# Post-implantation 3-T MRI in patients with an epicranial stimulation device: image quality and deep learning-based electrode reconstruction

**DOI:** 10.64898/2026.10.27.26366576

**Authors:** Niels Schwaderlapp, Antonia Barghoorn, Sotirios Kalousios, Matthias Dümpelmann, Eva Martinez-Lizana, Nicolas Zabler, Bernhard J. Steinhoff, Vera Dinkelacker, Maxim Zaitsev, Andreas Schulze-Bonhage

## Abstract

We evaluated structural and functional MRI at 3 T in five patients with an implanted EASEE focal cortex stimulation system and assessed MRI-only electrode reconstruction. Image quality was evaluated visually and by comparing implant-centered and control regions using structural GM/WM signal ratios, GE-EPI signal intensity, T2*, and relative off-resonance. A deep learning pipeline trained exclusively on synthetic data estimated electrode position from T1-weighted MPRAGE images. Structural and functional acquisitions were completed in all patients without scan-related adverse events. Structural images showed clear intracranial anatomy beneath the implant without visually apparent local signal loss or distortion. GE-EPI reference images and derived maps showed no consistent local disturbance corresponding to the electrode. GE-EPI signal was numerically lower in implant-centered regions, but no imaging metric differed significantly between regions (all p > 0.05). Electrode reconstructions were anatomically plausible in all patients, with patient-specific median central electrode-to-cortical gray-matter distances ranging from 9 to 20 mm. These findings demonstrate the feasibility of post-implantation structural and functional MRI and MRI-only estimation of electrode–cortex geometry, although localization accuracy remains unvalidated. This combination provides a basis for future studies integrating patient-specific electrode geometry, electric-field modeling, and functional connectivity analyses in focal cortex stimulation with the EASEE system.

**Plain Language Summary:** The EASEE system is an implant used to stimulate a specific brain region in people with focal epilepsy. In five patients, 3-T MRI provided clear images of brain anatomy and allowed functional brain scans despite the implant, while an artificial intelligence method estimated the electrode position directly from the MRI. This approach may help assess how the electrode is positioned relative to the underlying brain without requiring an additional CT scan.

## Introduction

Epicranial focal cortex stimulation (FCS) with the EASEE system (PRECISIS, Heidelberg, Germany) is an adjunctive neuromodulatory treatment for patients with drug-resistant focal epilepsy and a predominant epileptogenic region. The electrode is implanted epicranially over the individually defined target region, enabling focal transcranial stimulation of the epileptic focus without intracranial electrode placement. A prospective clinical trial demonstrated a sustained reduction in seizure frequency at 2-year follow-up, with favorable tolerability ^1,2^.

High-resolution structural MRI is central to the evaluation of focal epilepsy. The International League Against Epilepsy (ILAE) recommends high-resolution 3D T1-weighted and FLAIR imaging together with T2-weighted imaging as part of the HARNESS-MRI protocol ^3^. At 3 T, increased signal-to-noise ratio and image contrast support high-spatial-resolution imaging, while the higher field strength compared to 1.5 T increases sensitivity to susceptibility effects ^3,4^. Although the EASEE system is MR Conditional at 3 T under specified conditions, MR Conditional labeling addresses scanning safety rather than preservation of intracranial image quality. To our knowledge, clinical post-implantation 3-T MRI data in patients with an implanted EASEE system have not been reported.

For focal stimulation, the spatial relationship between the epicranial electrode and the underlying cortical anatomy may influence the magnitude and spatial distribution of the electric field delivered to the target region ^5,6^. For epicranial FCS, experimental work further indicates that, compared with transcutaneous stimulation, the shorter electrode-to-brain distance and the lack of skin impedance contribute to higher cortical voltage gradients ^7^. Accordingly, EASEE implantation often uses neuronavigation to align the electrode with the individually defined stimulation target ^8^. Post-implantation MRI-based visualization of the EASEE electrode has been proposed, but clinical results have not yet been reported ^9^.

In this study, we assessed structural and functional MRI image quality at 3 T in patients with an implanted EASEE system and developed an MRI-only approach to reconstruct the epicranial electrode and quantify its distance to cortical gray matter.

## Methods

### Participants and study design

Post-implantation MRI data were acquired as part of the prospective, single-center IMAGINE-STIM Upper Rhine study at the University Medical Center Freiburg. The parent study investigates multimodal measures of cerebral network changes in adults with drug-resistant focal epilepsy treated with the EASEE III system. The study was approved by the Ethics Committee of the University of Freiburg (24-1260-S1) and prospectively registered in the German Clinical Trials Register (DRKS00034611).

3-T MRI data from the first five participants with an implanted EASEE system for whom post-implantation MRI data were available were included.

### MRI acquisition

MRI was performed on a 3-T Siemens MAGNETOM Prisma scanner (Siemens Healthineers, Erlangen, Germany) using a 64-channel head coil. Structural imaging comprised sagittal 3D T1-weighted MPRAGE (1.0-mm isotropic resolution; TR/TE/TI = 2300/3.88/905 ms; flip angle = 9°, fat-suppressed), T2-weighted SPACE (1.0-mm isotropic; TR/TE = 3200/408 ms), and T2-weighted SPACE Dark Fluid (FLAIR; 1.0-mm isotropic; TR/TE/TI = 5000/386/1800 ms). Functional MRI (fMRI) was acquired using multi-echo gradient-echo EPI (3.0-mm isotropic; 48 slices; TR = 1000 ms; TE = 13.20/30.48/47.74 ms; flip angle = 60°; multiband factor = 4). Single-band reference images acquired as part of the fMRI protocol were used for susceptibility-sensitive image-quality assessment. Acquisition parameters are provided in Supplementary Methods 1.

All examinations followed the device-specific MR-conditional requirements of the implanted EASEE system ^10^.

### Image preprocessing and tissue segmentation

Structural images were defaced using pydeface ^11^ and coregistered to the T1-weighted MPRAGE using FLIRT (FSL, FMRIB Software Library, UK). Gray- and white-matter masks were generated using SimNIBS 4.6 ^5^.

### Quantitative image-quality assessment

The center coordinate of the reconstructed electrode mask defined a spherical implant-centered region of interest (ROI) with a radius of 35 mm, corresponding approximately to the spatial extent of the complete EASEE electrode. For structural images, the spherical ROI was intersected separately with the gray- and white-matter masks; for EPI-derived measures, it was intersected with the brain mask. Control ROIs were manually placed in homologous regions of the contralateral hemisphere (Supplementary Figure 1).

For the structural sequences, mean gray-matter (GM) and white-matter (WM) signal intensities were calculated separately and expressed as a GM/WM signal ratio. Mean GE-EPI signal within each ROI was normalized to the mean signal within the whole-brain EPI mask. Voxel-wise T2* maps were derived from the three echo magnitudes using a mono-exponential signal model and linear least-squares fitting of log-transformed signal versus echo time. Relative off-resonance maps were calculated from the three phase images using spatial phase unwrapping followed by voxel-wise linear fitting of phase versus echo time. The frequency maps were referenced to the whole-brain median and therefore represent relative rather than absolute off-resonance.

### MRI-only electrode reconstruction

Electrode position was estimated from post-implantation fat-suppressed T1-weighted MPRAGE using a deep learning pipeline trained exclusively on synthetic implant examples generated from healthy-participant MRI. The pipeline localized implant-associated signal voids, segmented the electrode contacts, and fitted a template of the known electrode geometry. Details in Supplementary Methods 2 and the GitHub repository ^12^.

### Central electrode-to-cortical gray-matter distance

Distances to cortical gray matter were assessed only for the central electrode because of its cathodal role in DC-like stimulation. A 20-mm-diameter disk, matching its outer diameter, was centered on and aligned with the reconstructed electrode. Sampling points were placed on a 0.5-mm grid within the disk. Distances were measured perpendicular to the electrode plane toward the cortex to the first cortical gray-matter voxel in the SimNIBS segmentation. Patient-specific distributions were summarized by median, mean, minimum, and maximum and displayed as violin plots showing spatial variation beneath the electrode.

### Statistical analysis

Imaging metrics within implant-centered and contralateral control ROIs were summarized descriptively. Two-sided paired *t*-tests compared implant-centered and control regions for each of the six imaging metrics. Statistical significance was defined as α = 0.05. No adjustment for multiple comparisons was applied because the analyses were exploratory. Given the small sample size (*n* = 5), *p*-values were interpreted descriptively rather than as confirmatory evidence.

## Results

Post-implantation 3-T MRI, including structural and functional acquisitions, was successfully completed in all patients. No scan-related adverse events or clinically apparent device-related problems were observed.

Across all five patients, structural T1-weighted, T2-weighted, and FLAIR images showed clear visualization of intracranial anatomy beneath the implant, without visually apparent local signal loss or distortion (Figure 1A–C). Likewise, GE-EPI reference images acquired as part of the fMRI protocol, T2* maps, and relative off-resonance maps showed no consistent local signal disturbance spatially corresponding to the overlying electrode (Figure 1D–F). Images from all patients are provided in Supplementary Figure 2.

**Figure 1.**
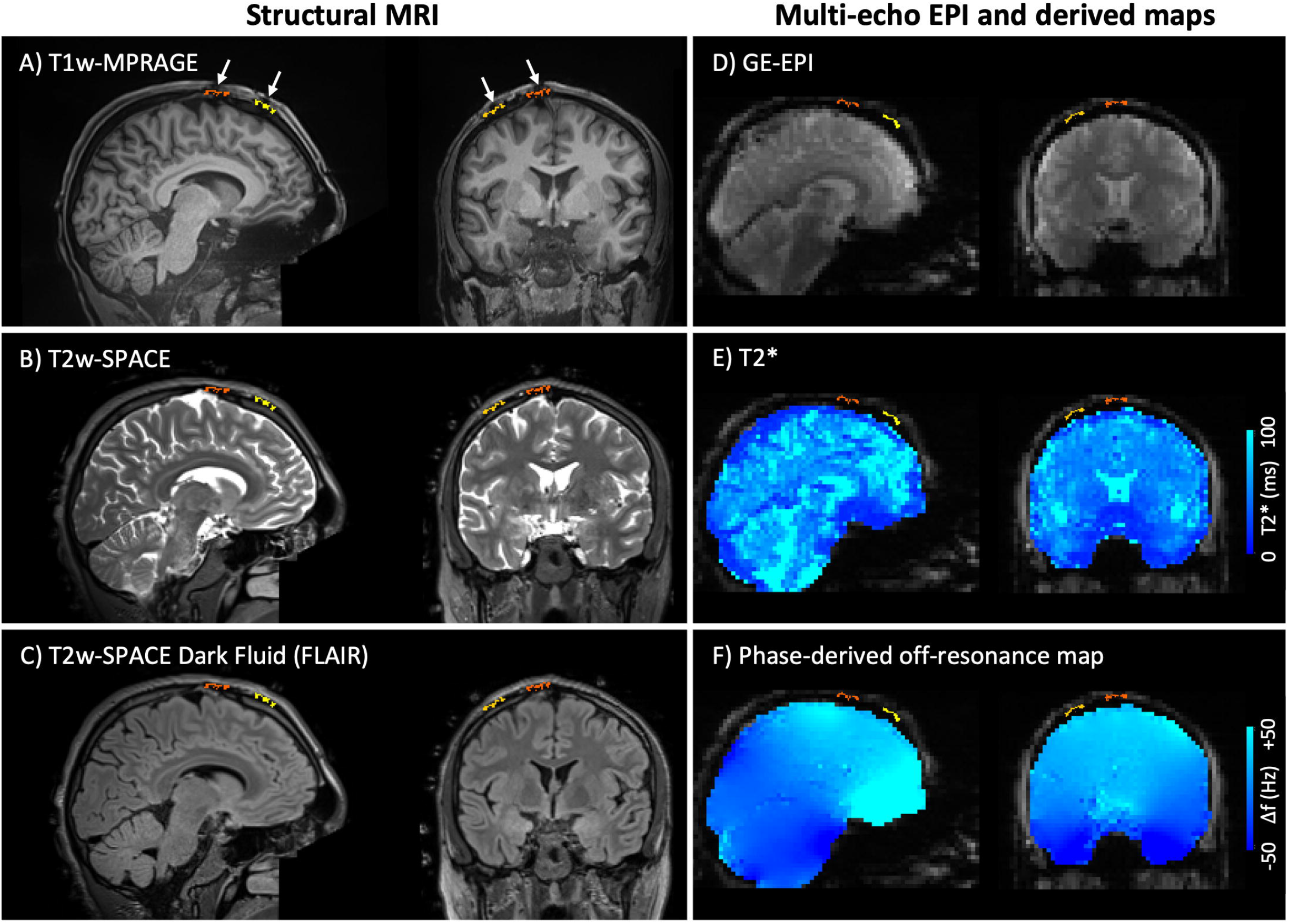
Representative post-implantation 3-T MRI in a patient with an implanted EASEE system. Matched sagittal and coronal views through the brain region underlying the epicranial electrode are shown. Structural imaging comprises **(A)** fat-suppressed T1-weighted MPRAGE, **(B)** T2-weighted SPACE, and **(C)** T2-weighted SPACE Dark Fluid (FLAIR). Susceptibility-sensitive imaging comprises **(D)** gradient-echo EPI magnitude images at the first echo (TE = 13.2 ms), **(E)** voxel-wise T2* maps, and **(F)** phase-derived relative off-resonance maps. Yellow/orange overlays indicate the MRI-based reconstructed electrode position; arrows in **(A)** indicate electrode-associated extracranial signal voids. In this representative example, these extracranial signal voids are most noticeable on the fat-suppressed T1-weighted image and are only minimally apparent on the T2-weighted and FLAIR images. Intracranial anatomy beneath the electrode remains clearly visualized on the structural images, without visually apparent local signal loss or distortion. No corresponding local intracranial signal disturbance is apparent on the GE-EPI magnitude, T2*, or relative off-resonance images.

Quantitative within-subject comparisons were consistent with the qualitative imaging findings (Figure 2). Gray-to-white matter contrast showed no consistent reduction in implant-centered regions across the structural acquisitions. GE-EPI signal intensity showed a numerical reduction in implant-centered regions at the group level, whereas T2* and relative off-resonance showed no consistent directional differences between implant-centered and contralateral control regions. None of the six paired comparisons reached statistical significance (all p > 0.05).

**Figure 2.**
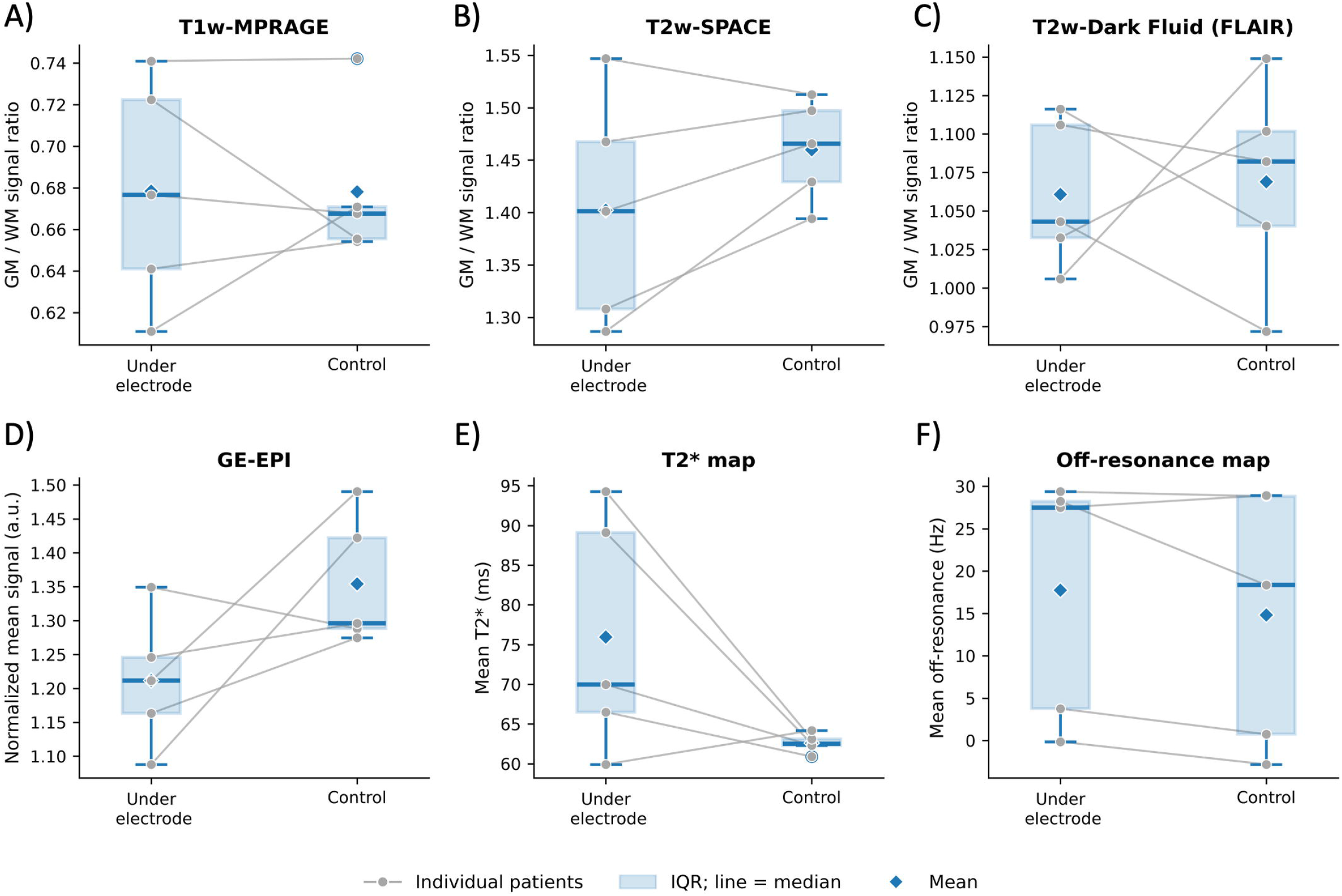
Quantitative comparison of MRI-derived image-quality metrics beneath the EASEE electrode and in contralateral control regions. Paired within-subject measurements from five patients are shown for implant-centered regions and their homologous contralateral control regions. **(A–C)** Gray-matter/white-matter (GM/WM) signal-intensity ratios for T1-weighted MPRAGE, T2-weighted SPACE, and T2-weighted SPACE Dark Fluid (FLAIR), respectively. **(D)** Mean gradient-echo EPI signal intensity normalized to the whole-brain EPI mean. **(E)** Mean T2* within the respective regions of interest, derived from the three-echo magnitude data. **(F)** Mean phase-derived relative off-resonance. Gray circles represent individual patients, with connecting lines indicating paired measurements. Boxes indicate the interquartile range, horizontal lines the median, and diamonds the mean. Two-sided paired *t*-tests showed no statistically significant differences between implant-centered and control regions for any metric (**A**, *p* = 0.988; **B**, *p* = 0.121; **C**, *p* = 0.856; **D**, *p* = 0.122; **E**, *p* = 0.117; **F**, *p* = 0.199).

A deep learning-based reconstruction model trained exclusively on synthetic data (Figure 3A– C) yielded anatomically plausible EASEE electrode reconstructions from post-implantation T1w-MPRAGE (Figure 3D, E). Reconstructions from all patients are shown in Supplementary Figure 3. Quantification of central electrode-to-cortical gray-matter distance demonstrated interindividual variability across the five patients, with median distances ranging from 9 to 20 mm and minimum distances from 4.5 to 11.5 mm (Figure 3F).

**Figure 3.**
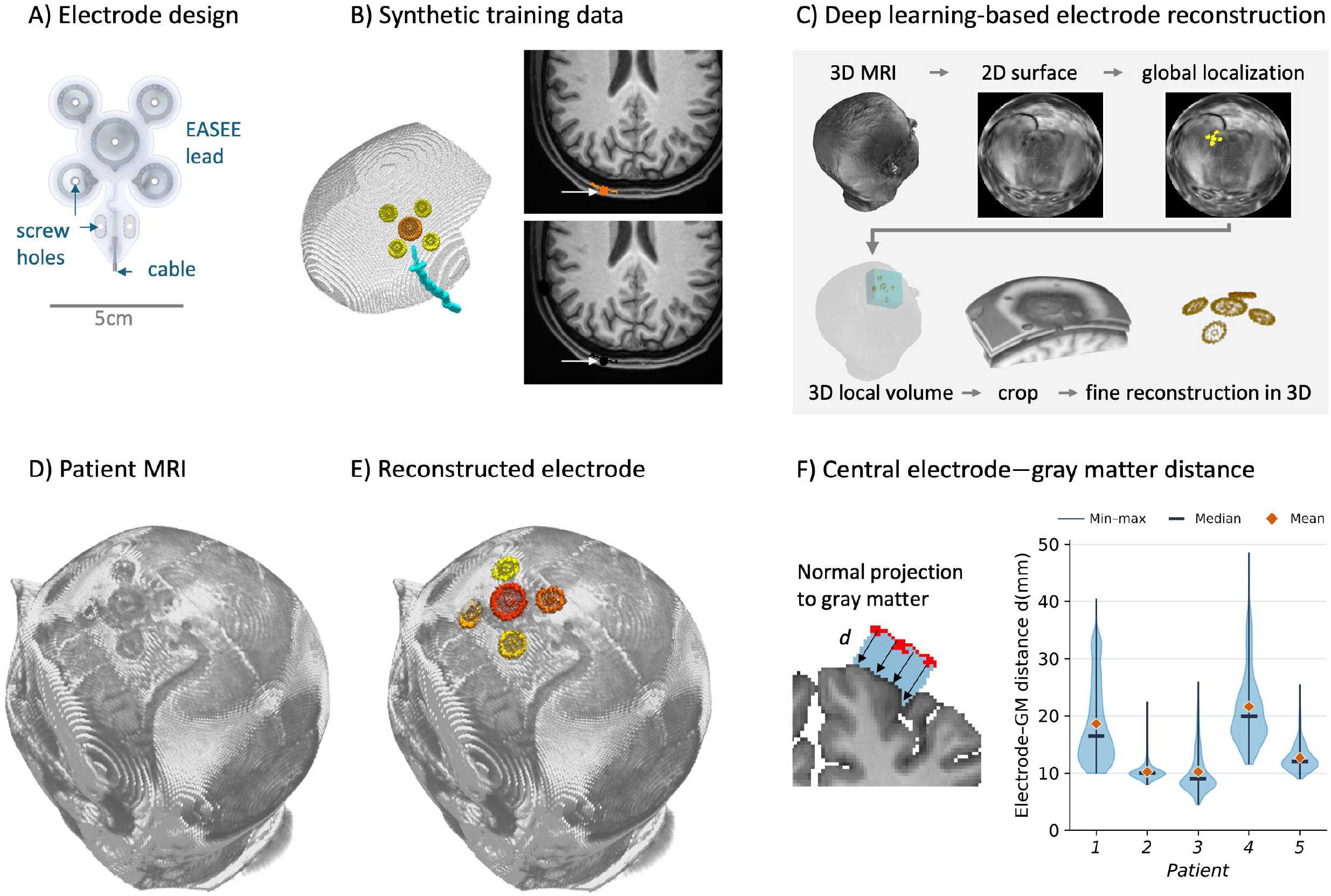
MRI-only reconstruction of the EASEE electrode and patient-specific electrode-to-cortical gray-matter geometry. **(A)** Geometry of the EASEE lead, including the central and four peripheral electrode contacts, fixation holes, and connecting cable. **(B)** Generation of synthetic training data. EASEE implants were simulated at anatomically plausible positions on MRI-derived skull surfaces of healthy participants, including the electrode, cable, and corresponding implant-associated MR signal alterations. **(C)** Two-stage deep learning-based reconstruction workflow. Two-dimensional projections at multiple depths relative to the head surface were used for global electrode localization by a 2D U-Net. A local three-dimensional MRI volume centered on the predicted position was subsequently processed by a 3D U-Net to reconstruct the electrode geometry. **(D)** Representative post-implantation T1-weighted MRI showing the subtle extracranial implant signature, visualized after clipping approximately 10 mm of the overlying superficial tissue to expose the cranial surface. **(E)** MRI-only electrode reconstruction overlaid on the corresponding patient-specific anatomy. **(F)** Patient-specific electrode-to-cortical gray-matter distance across a disk aligned with the central electrode plane. Violin plots show the distribution of point-wise distances for each patient; horizontal bars indicate the median, vertical lines the minimum-to-maximum range, and diamonds the mean.

## Discussion

The principal finding of this study is that structural 3-T MRI retained clear visualization of intracranial anatomy and GM/WM signal-intensity ratio beneath the implanted EASEE electrode, while its extracranial MRI signature was sufficiently distinct to permit MRI-only reconstruction relative to the individual cortical anatomy.

Preserved structural image quality is relevant to focal epilepsy because detecting subtle cortical abnormalities requires clear delineation of cortical anatomy and gray–white matter boundaries ^3^. Studies of VNS, DBS, and the responsive neurostimulation system illustrate that MRI feasibility does not ensure preserved image quality, although differences in hardware and acquisition protocols limit direct comparison with the EASEE system ^13–16^. In our study, structural images showed no visually apparent local signal loss or distortion beneath the epicranial electrode, and GM/WM signal-intensity ratios showed no consistent reduction relative to contralateral control regions.

Beyond structural imaging, the successful acquisition of fMRI data demonstrates the feasibility of functional MRI in patients with implanted EASEE systems. GE-EPI, the basis of conventional BOLD-fMRI, is particularly sensitive to B_0_ inhomogeneity and off-resonance, which can cause signal loss and geometric distortion ^17^. In this cohort, GE-EPI signal intensity was numerically lower in implant-centered regions, but the difference was not statistically significant and not accompanied by systematic changes in T2* or relative off-resonance. The data therefore do not provide evidence for a consistent local susceptibility-related effect, and the GE-EPI observation should remain descriptive rather than being attributed to a specific implant-related mechanism.

MRI-only electrode reconstruction addresses a second aspect specific to FCS: characterization of the implant’s spatial relationship to individual anatomy. EASEE implantation is spatially targeted, and alignment of the central cathodal contact with the intended stimulation point is considered important for stimulation delivery ^7,8^. The electrode reconstructions enabled estimation of electrode-to-cortical gray-matter distances, with patient-specific medians ranging from 9 to 20 mm. Beyond distance, anatomical features such as local skull thickness, cerebrospinal fluid volume, and cortical morphology could also influence electric field distribution ^18^. Such characterization may be particularly relevant when the targeted region contains structural abnormalities, including focal tissue loss or altered cortical thickness associated with the underlying epileptogenic pathology ^19^. The present distance analysis therefore illustrates a broader approach to quantifying patient-specific implant–anatomy relationships. Whether these anatomical measures explain differences in stimulation fields or clinical response requires further investigation combining MRI-derived geometry, individualized electric-field modeling, and clinical outcomes.

## Limitations

The main limitation is the absence of an independent reference standard for electrode position, such as postoperative CT; therefore, MRI-only reconstruction demonstrates feasibility but does not establish localization accuracy. The cohort comprised five patients, limiting the assessment of subtle sequence-specific imaging effects. In addition, the present study was not designed to determine whether MRI-derived geometric parameters are related to stimulation efficacy, which will require larger prospective cohorts.

### Clinical Relevance and Future Directions

This study demonstrates that implantation of the EASEE device for FCS preserves the ability of post-implantation 3-T MRI to provide both high-resolution structural and functional brain imaging. Furthermore, MRI-only electrode reconstruction enables the implanted electrode to be related directly to the individual cortical anatomy without requiring radiation exposure by performing CT for reconstruction. This spatial information could inform patient-specific electric-field modeling and is particularly relevant for epicranial stimulation, as individual tissue geometry influences cortical field strength.

The ability to acquire fMRI after implantation also creates opportunities to investigate changes in functional connectivity over the course of treatment. Combining these approaches will allow future studies to determine whether anatomical and functional imaging markers are associated with stimulation efficacy and whether optimization of electrode positioning relative to the underlying epileptogenic brain region can ultimately contribute to treatment optimization or response prediction.

## Data Availability

The code for localizing and reconstructing the EASEE electrodes is available in the corresponding GitHub repository. The patients' MRI data cannot be made publicly available because characteristic electrode patterns prevent sufficient anonymization.

https://github.com/N-Schwaderlapp/EASEE_localization

## Acknowledgements

This work was co-financed by the European Union (Interreg Project IMAGINE-STIM).

## Ethical Publication Statement

“We confirm that we have read the Journal’s position on issues involved in ethical publication and affirm that this report is consistent with those guidelines.”

## Statement on Data Availability

The code for localizing and reconstructing the EASEE electrodes is available in the corresponding GitHub repository ^12^. The patients’ MRI data cannot be made publicly available because characteristic electrode patterns prevent sufficient anonymization.

