## Supplementary Figure 1 for "Post-implantation 3-T MRI in patients with an epicranial stimulation device: image quality and deep learning-based electrode reconstruction"

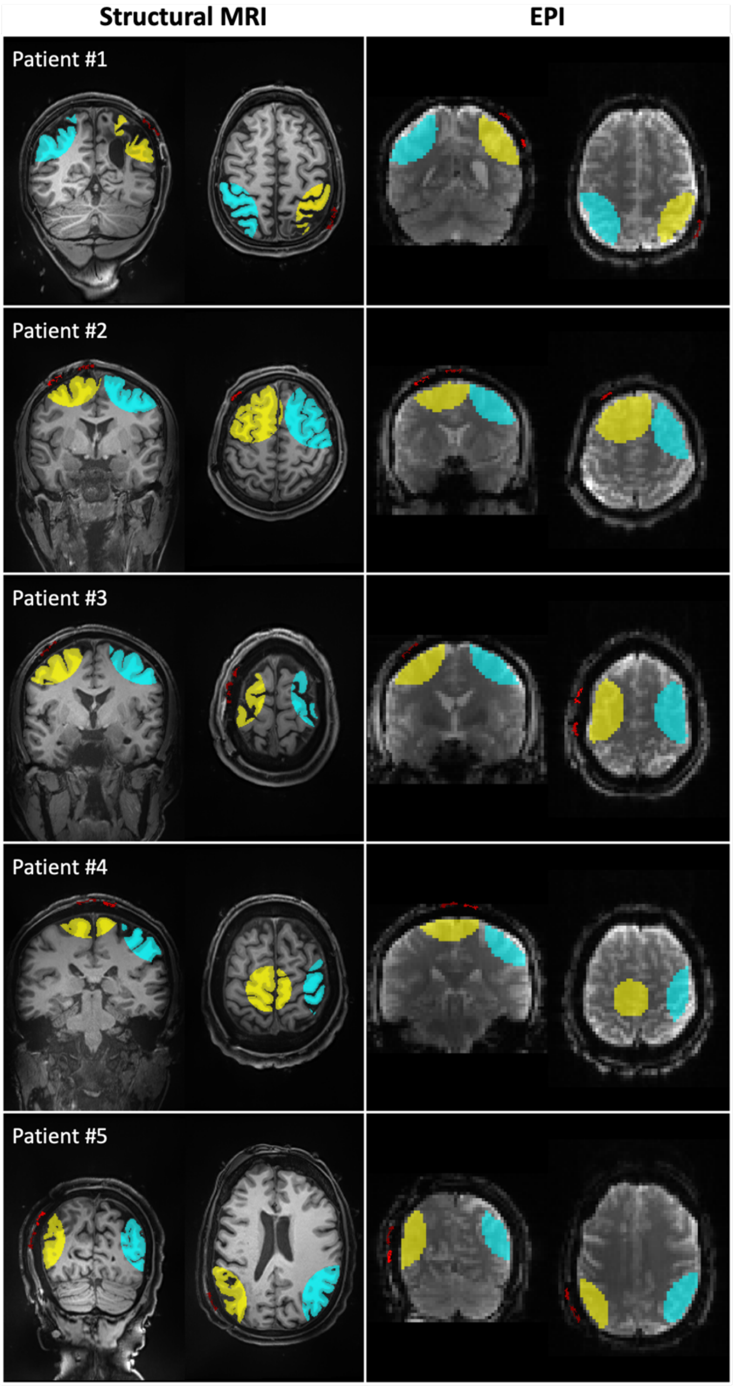


**Supplementary Figure 1. Implant-centered and contralateral control regions of interest used for quantitative MRI analysis.**
Representative coronal and axial views are shown for all five patients (top to bottom). The left column shows regions of interest (ROIs) used for the structural MRI analyses, and the right column shows the corresponding ROIs used for gradient-echo EPI analyses. Yellow ROIs indicate the implant-centered region beneath the EASEE electrode, whereas cyan ROIs indicate manually selected control regions in the contralateral hemisphere. Control ROIs were positioned at anatomically corresponding locations to match tissue architecture and imaging depth as closely as possible. Red overlays indicate the reconstructed position of the EASEE electrode. These ROIs were used for the paired within-subject comparisons of MRI-derived image-quality metrics shown in Figure 2.
