## Supplementary figures and images for "Post-implantation 3-T MRI in patients with an epicranial stimulation device: image quality and deep learning-based electrode reconstruction"

### Supplementary Figure 2

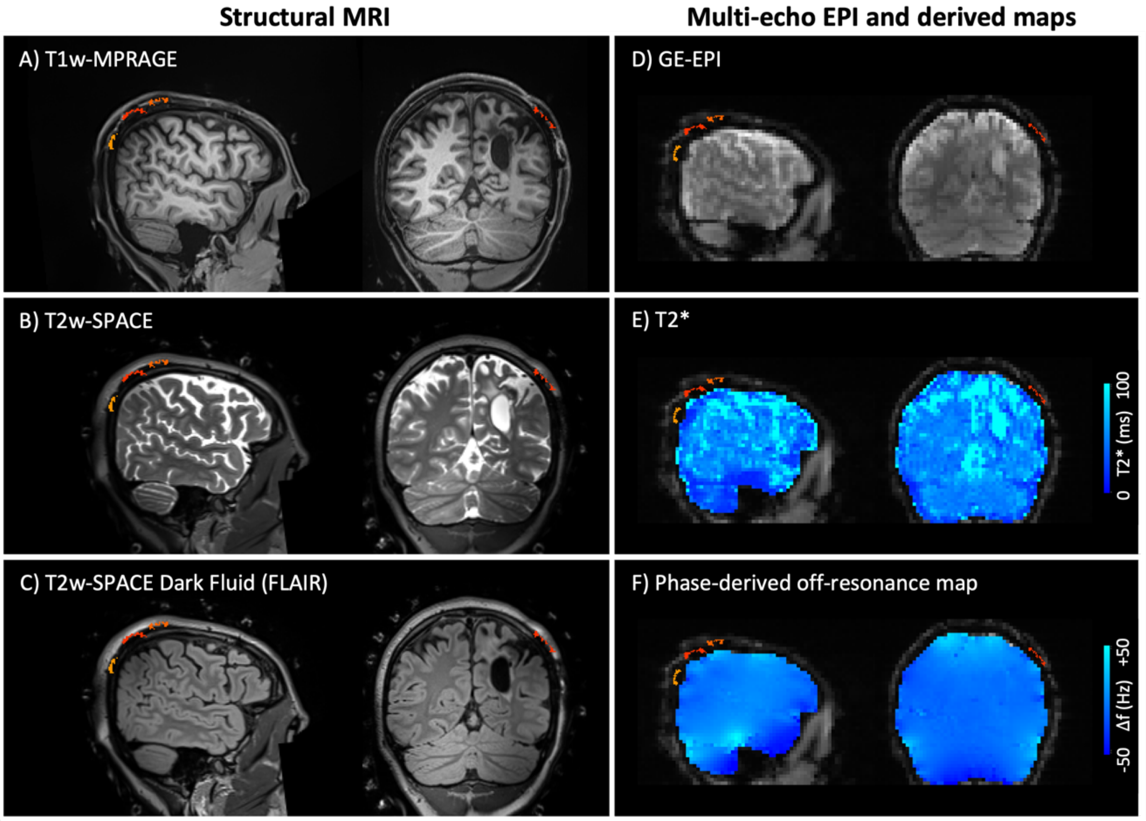


Patient #1

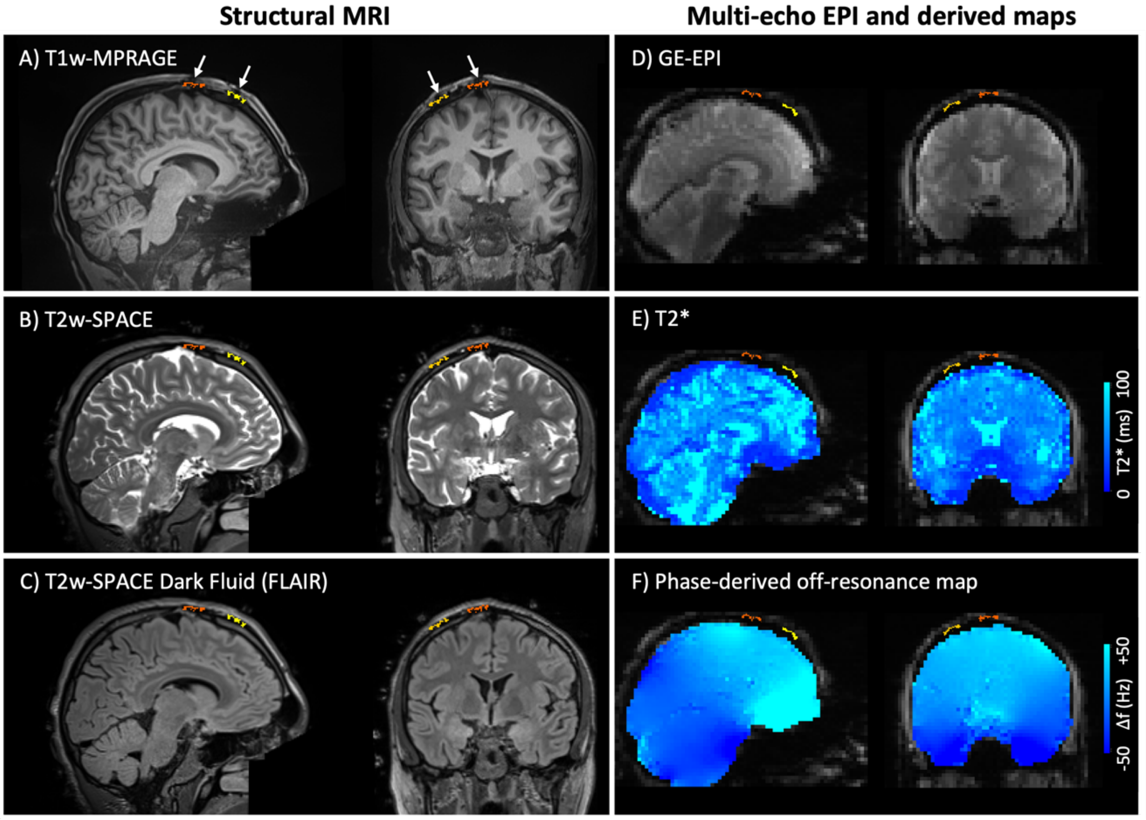


Patient #2

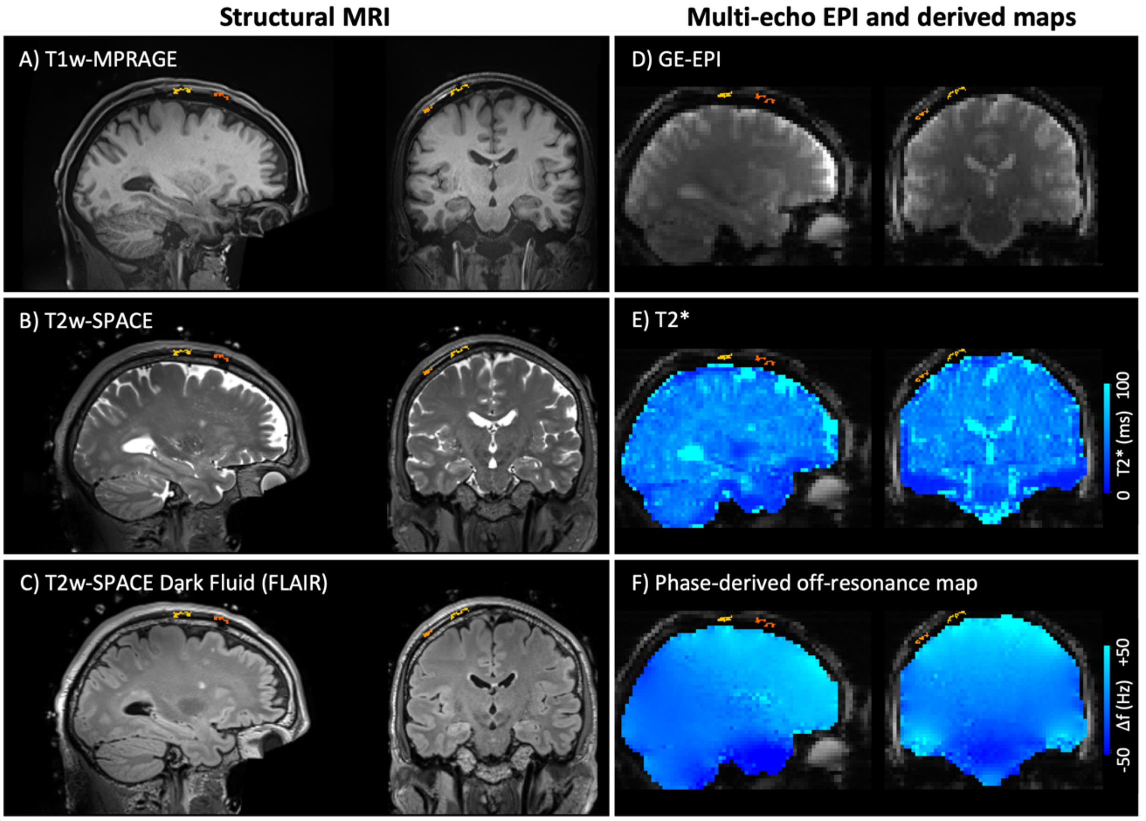


Patient #3

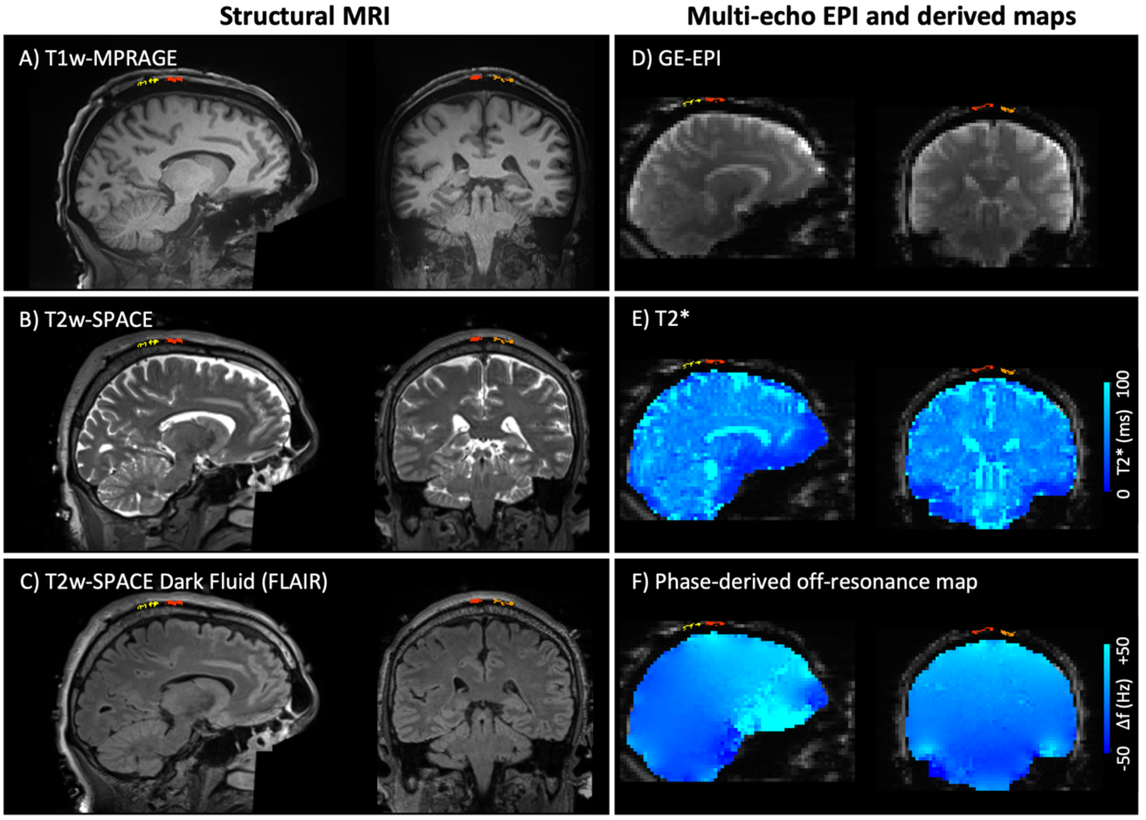


Patient #4

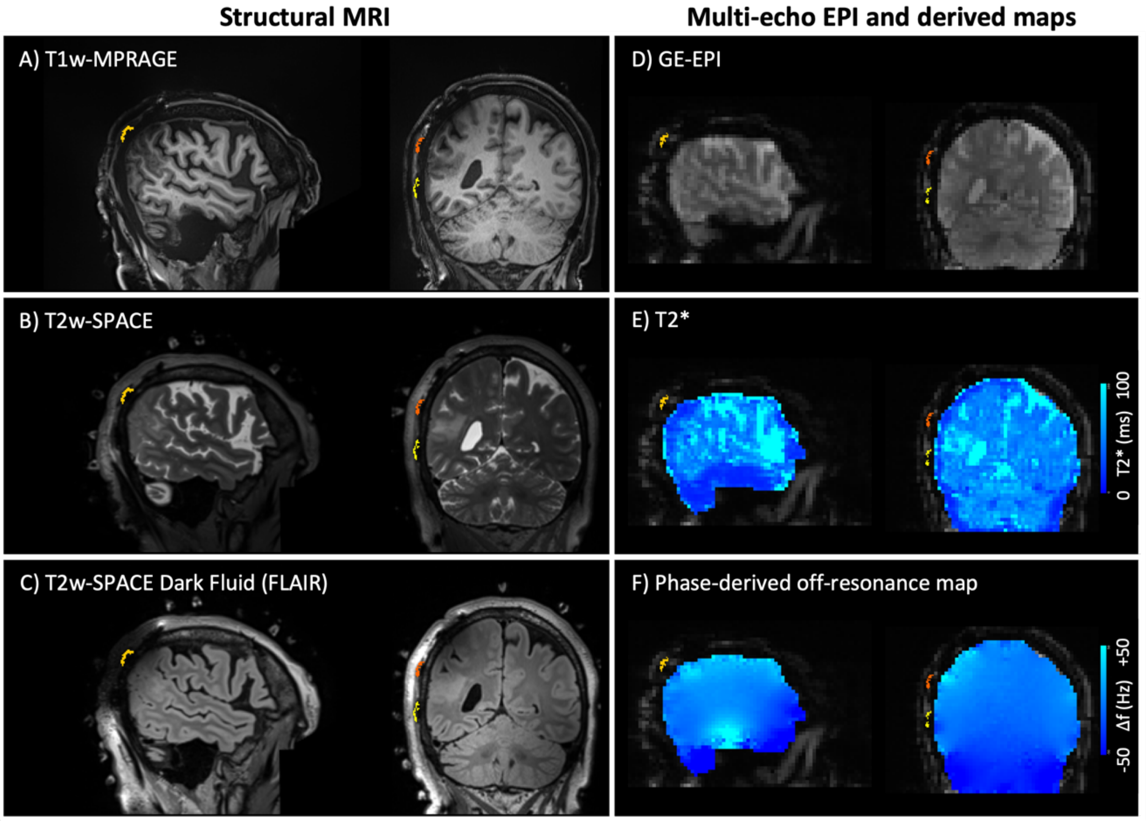
