## Supplementary Figure 3 for "Post-implantation 3-T MRI in patients with an epicranial stimulation device: image quality and deep learning-based electrode reconstruction"

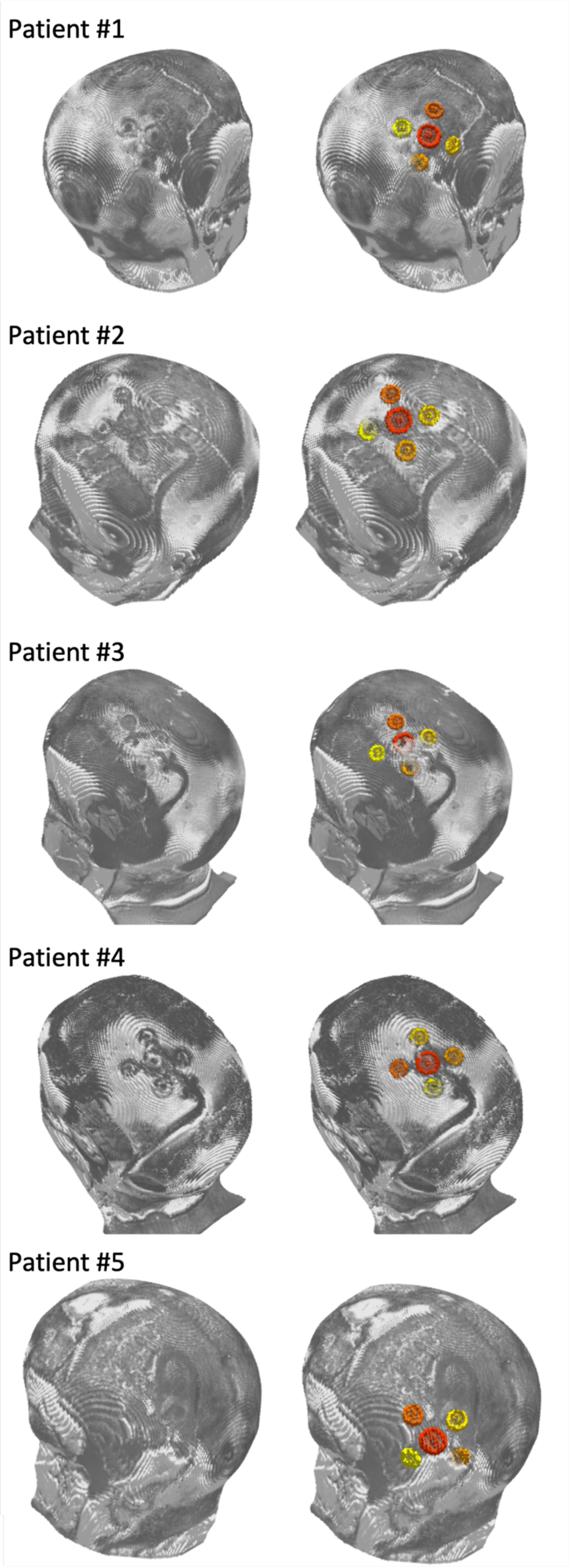


**Supplementary Figure 3. MRI-only reconstruction of the EASEE electrode.**

For all five patients (top to bottom), post-implantation T1-weighted MRI showing the subtle extracranial implant signature, visualized after clipping approximately 10 mm of the overlying superficial tissue to expose the cranial surface (left column). MRI-only electrode reconstruction overlaid on the corresponding patient-specific anatomy (right column).
