## Supplementary Methods 1 for "Post-implantation 3-T MRI in patients with an epicranial stimulation device: image quality and deep learning-based electrode reconstruction"

Supplementary: MRI acquisition protocols

1. T1-weighted MPRAGE
2. T2-weighted SPACE
3. T2-weigthed SPACE Dark Fluid (FLAIR)
4. Gradient Echo EPI

### \\MedPhys\ABI\Schwaderlapp\EASEE_Studie_v2\T1w_MPRAGE_1mm

TA: 4:44 min Coil Selection: Manual Voxel Size: 1.0×1.0×1.0 mm³ Acc:: 2 Rel. SNR: 1.00

#### Properties

| | |
|---|---|
| Start measurement without further preparation | On |
| Wait for User to Start | On |
| Start measurements | Single Measurement |
| Prio Recon | Off |
| Auto Open Inline Display | Off |
| Auto Close Inline Display | Off |
| Load Images to MR View&GO | On |
| Auto Store Images | On |
| Disable auto transfer to PACS | Off |
| Load Images to Stamp Segments | Off |
| Load Images to Graphic Segments | Off |
| Graphic segment | Default |
| Inline Movie | Off |

#### Routine

| | |
|---|---|
| Slab Group | 1 |
| Slabs | 1 |
| Distance Factor | 50 % |
| Position | L0.0 P0.0 F9.5 mm |
| Orientation | Sagittal |
| Phase Encoding Dir. | A >> P |
| Slices per Slab | 176 |
| Phase Oversampling | 0 % |
| Slice Oversampling | 0.0 % |
| FOV Read | 256 mm |
| FOV Phase | 100.0 % |
| Slice Thickness | 1.00 mm |
| TR | 2300.0 ms |
| TE | 3.88 ms |
| Averages | 1 |
| Concatenations | 1 |
| AutoAlign | Head > Brain |
| Coil Elements | HC1-7;NC1,2 |

#### Contrast - Common

| | |
|---|---|
| TR | 2300.0 ms |
| TE | 3.88 ms |
| Magn. Preparation | Non-sel. IR |
| TI | 905 ms |
| Flip Angle | 9 deg |
| Fat-Water Contrast | Water Excitation |
| Dark Blood | Off |
| Contrasts | 1 |
| Reconstruction | Magnitude |

#### Contrast - Dynamic

| | |
|---|---|
| Dynamic Mode | Standard |
| Measurements | 1 |
| Multiple Series | Each Measurement |
| Reordering | Linear |

#### Resolution - Common

| | |
|---|---|
| FOV Read | 256 mm |
| FOV Phase | 100.0 % |
| Slice Thickness | 1.00 mm |
| Base Resolution | 256 |
| Phase Resolution | 100 % |
| Slice Resolution | 100 % |
| Interpolation | Off |

#### Resolution - Acceleration

| | |
|---|---|
| Acceleration Mode | GRAPPA |
| Reference Scans | Integrated |
| Acceleration Factor PE | 2 |
| Reference Lines PE | 24 |
| Acceleration Factor 3D | 1 |
| Phase Partial Fourier | 7/8 |
| Slice Partial Fourier | Off |
| Asymmetric Echo | Allowed |
| Elliptical Scanning | Off |

#### Resolution - Filter

| | |
|---|---|
| Raw Filter | Off |
| Elliptical Filter | Off |
| Distortion Correction | 2D |
| Normalize | Prescan |
| Image Filter | On |

#### Geometry - Common

| | |
|---|---|
| Slab Group | 1 |
| Slabs | 1 |
| Distance Factor | 50 % |
| Position | L0.0 P0.0 F9.5 mm |
| Orientation | Sagittal |
| Phase Encoding Dir. | A >> P |
| Slices per Slab | 176 |
| Phase Oversampling | 0 % |
| Slice Oversampling | 0.0 % |
| FOV Read | 256 mm |
| FOV Phase | 100.0 % |
| Slice Thickness | 1.00 mm |
| TR | 2300.0 ms |
| Multi-Slice Mode | Single Shot |
| Series | Ascending |
| Concatenations | 1 |

#### Geometry - AutoAlign

| | |
|---|---|
| Slab Group | 1 |
| Position | L0.0 P0.0 F9.5 mm |
| Orientation | Sagittal |
| Phase Encoding Dir. | A >> P |
| AutoAlign | Head > Brain |

#### Geometry - AutoAlign

| | |
|---|---|
| Initial Position | Isocenter |
| R | 0.0 mm |
| P | 0.0 mm |
| F | 0.0 mm |
| Initial Orientation | Sagittal |
| Initial Rotation | 0.00 deg |

#### Geometry - Navigator

#### Geometry - Tim Planning Suite

| | |
|---|---|
| Set-n-Go Protocol | Off |
| Table Position | 10 mm |
| Table Position | F |
| Inline Composing | Off |

#### System - Miscellaneous

| | |
|---|---|
| Coil Selection | Manual |
| Radial Sorting | Off |
| MSMA | S - C - T |
| Sagittal | R >> L |
| Coronal | A >> P |
| Transversal | F >> H |
| Coil Combination | Sum of Squares |
| Matrix Optimization | Off |

#### System - Adjustments

| | |
|---|---|
| Adjustment Strategy | Standard |
| B0 Shim | Standard |
| B1 Shim | TrueForm |
| Adjustment Tolerance | Auto |
| Adjust with Body Coil | Off |
| Confirm Frequency | Never |
| Assume Silicone | Off |

#### System - Adjust Volume

| | |
|---|---|
| Position | L0.0 P0.0 F9.5 mm |
| Orientation | Sagittal |
| Rotation | 0.00 deg |
| A >> P | 256 mm |
| F >> H | 256 mm |
| R >> L | 176 mm |
| Reset | Off |

#### System - pTx

| | |
|---|---|
| B1 Shim | TrueForm |
| Excitation | Slab-sel. |

#### System - Tx/Rx

| | |
|---|---|
| Frequency 1H | 123.194104 MHz |
| ? Ref. Amplitude 1H | 0.000 V |
| Reset | Off |
| Correction Factor | 1.00 |
| Image Scaling | 1.000 |

#### Physio - Signal

| | |
|---|---|
| 1st Signal/Mode | None |
| TR | 2300.0 ms |
| Concatenations | 1 |

#### Physio - Cardiac

| | |
|---|---|
| Fat-Water Contrast | Water Excitation |
| Magn. Preparation | Non-sel. IR |
| TI | 905 ms |
| Dark Blood | Off |
| FOV Read | 256 mm |
| FOV Phase | 100.0 % |
| Phase Resolution | 100 % |

#### Physio - PACE

| | |
|---|---|
| Resp. Control | Off |
| Concatenations | 1 |

#### Inline - Subtraction

| | |
|---|---|
| Subtract | Off |
| Measurements | 1 |
| StdDev | Off |
| Save Original Images | On |

#### Inline - MIP

| | |
|---|---|
| MIP Sag | Off |
| MIP Cor | Off |
| MIP Tra | Off |
| MIP Time | Off |
| Radial MIP | Off |
| Save Original Images | On |
| MPR Sag | Off |
| MPR Cor | Off |
| MPR Tra | Off |

#### Inline - Composing

| | |
|---|---|
| Inline Composing | Off |

#### Inline - MapIt

| | |
|---|---|
| MapIt | None |
| Flip Angle | 9 deg |
| Measurements | 1 |
| Contrasts | 1 |
| TE | 3.88 ms |
| TR | 2300.0 ms |
| Save Original Images | On |

#### Sequence - Part 1

| | |
|---|---|
| Sequence Name | tfl |
| Dimension | 3D |
| Excitation | Slab-sel. |
| RF Pulse Type | Fast |
| Gradient Mode | Whisper |
| Flow Compensation | None |
| Reordering | Linear |

#### Sequence - Part 1

| | |
|---|---|
| Bandwidth | 240 Hz/Px |
| Echo Spacing | 10.04 ms |
| Asymmetric Echo | Allowed |
| Turbo Factor | 176 |

#### Sequence - Part 2

| | |
|---|---|
| Introduction | On |
| RF Spoiling | On |
| Incr. Gradient Spoiling | On |

#### Sequence - Assistant

| | |
|---|---|
| SAR Assistant | Off |

| \\MedPhys\ABI\Schwaderlapp\EASEE_Studie_v2\T2w_space_sag_1mm_iso |
|---|
| TA: 4:11 min Coil Selection: Manual Voxel Size: 1.0×1.0×1.0 mm³ Acc:: 2 Rel. SNR: 1.00 |

#### Properties

| | |
|---|---|
| Start measurement without further preparation | Off |
| Wait for User to Start | On |
| Start measurements | Single Measurement |
| Prio Recon | Off |
| Auto Open Inline Display | Off |
| Auto Close Inline Display | Off |
| Load Images to MR View&GO | On |
| Auto Store Images | On |
| Disable auto transfer to PACS | Off |
| Load Images to Stamp Segments | On |
| Load Images to Graphic Segments | Off |
| Graphic segment | Default |
| Inline Movie | Off |

#### Routine

| | |
|---|---|
| Slab Group | 1 |
| Slabs | 1 |
| Position | Isocenter |
| Orientation | Sagittal |
| Phase Encoding Dir. | A >> P |
| Slices per Slab | 176 |
| Phase Oversampling | 0 % |
| Slice Oversampling | 0.0 % |
| FOV Read | 256 mm |
| FOV Phase | 100.0 % |
| Slice Thickness | 1.00 mm |
| TR | 3200.0 ms |
| TE | 408.00 ms |
| Averages | 1.0 |
| Concatenations | 1 |
| AutoAlign | Head > Brain |
| Coil Elements | HC1-7;NC1,2 |

#### Contrast - Common

| | |
|---|---|
| TR | 3200.0 ms |
| TE | 408.00 ms |
| MTC | Off |
| Magn. Preparation | None |
| Flip Angle Mode | T2 Var |
| Fat-Water Contrast | Standard |
| Dark Blood | Off |
| Blood Suppression | Off |
| Wrap-up Magn. | Restore |
| Reconstruction | Magnitude |

#### Contrast - Dynamic

| | |
|---|---|
| Dynamic Mode | Standard |
| Measurements | 1 |
| Multiple Series | Each Measurement |
| Reordering | Linear |

#### Resolution - Common

| | |
|---|---|
| FOV Read | 256 mm |
| FOV Phase | 100.0 % |
| Slice Thickness | 1.00 mm |
| Base Resolution | 256 |
| Phase Resolution | 100 % |
| Slice Resolution | 100 % |
| Interpolation | Off |

#### Resolution - Acceleration

| | |
|---|---|
| Acceleration Mode | GRAPPA |
| Total Factor | 2 |
| Reference Scans | Integrated |
| Acceleration Factor PE | 2 |
| Reference Lines PE | 24 |
| Acceleration Factor 3D | 1 |
| Phase Partial Fourier | Allowed |
| Slice Partial Fourier | 7/8 |
| Elliptical Scanning | Off |

#### Resolution - Filter

| | |
|---|---|
| Raw Filter | On |
| Elliptical Filter | Off |
| Distortion Correction | 2D |
| Normalize | Prescan |
| Image Filter | Off |

#### Geometry - Common

| | |
|---|---|
| Slab Group | 1 |
| Slabs | 1 |
| Position | Isocenter |
| Orientation | Sagittal |
| Phase Encoding Dir. | A >> P |
| Slices per Slab | 176 |
| Phase Oversampling | 0 % |
| Slice Oversampling | 0.0 % |
| FOV Read | 256 mm |
| FOV Phase | 100.0 % |
| Slice Thickness | 1.00 mm |
| TR | 3200.0 ms |
| Concatenations | 1 |

#### Geometry - AutoAlign

| | |
|---|---|
| Slab Group | 1 |
| Position | Isocenter |
| Orientation | Sagittal |
| Phase Encoding Dir. | A >> P |
| AutoAlign | Head > Brain |
| Initial Position | Isocenter |
| L | 0.0 mm |
| P | 0.0 mm |

#### Geometry - AutoAlign

| | |
|---|---|
| H | 0.0 mm |
| Initial Orientation | Sagittal |
| Initial Rotation | -0.01 deg |

#### Geometry - Navigator

#### Geometry - Saturation

| | |
|---|---|
| Special Saturation | None |

#### Geometry - Tim Planning Suite

| | |
|---|---|
| Set-n-Go Protocol | Off |
| Table Position | 0 mm |
| Table Position | H |
| Inline Composing | Off |

#### System - Miscellaneous

| | |
|---|---|
| Coil Selection | Manual |
| Radial Sorting | Off |
| MSMA | S - C - T |
| Sagittal | R >> L |
| Coronal | A >> P |
| Transversal | F >> H |
| Coil Combination | Adaptive Combine |
| Matrix Optimization | Off |

#### System - Adjustments

| | |
|---|---|
| Adjustment Strategy | Standard |
| B0 Shim | Standard |
| B1 Shim | TrueForm |
| Adjustment Tolerance | Auto |
| Adjust with Body Coil | Off |
| Confirm Frequency | Never |
| Assume Silicone | Off |

#### System - Adjust Volume

| | |
|---|---|
| Position | Isocenter |
| Orientation | Sagittal |
| Rotation | -0.01 deg |
| A >> P | 256 mm |
| F >> H | 256 mm |
| R >> L | 176 mm |
| Reset | Off |

#### System - pTx

| | |
|---|---|
| B1 Shim | TrueForm |
| Excitation | Non-sel. |

#### System - Tx/Rx

| | |
|---|---|
| Frequency 1H | 123.194104 MHz |
| ? Ref. Amplitude 1H | 0.000 V |
| Reset | Off |
| Correction Factor | 1.00 |
| Image Scaling | 1.000 |
| Gain | High |

#### Physio - Signal

| | |
|---|---|
| 1st Signal/Mode | None |
| Trigger Delay | 0 ms |
| TR | 3200.0 ms |
| Concatenations | 1 |

#### Physio - Cardiac

| | |
|---|---|
| Fat-Water Contrast | Standard |
| Magn. Preparation | None |
| Dark Blood | Off |
| FOV Read | 256 mm |
| FOV Phase | 100.0 % |
| Phase Resolution | 100 % |

#### Physio - PACE

| | |
|---|---|
| Resp. Control | Off |
| Concatenations | 1 |

#### Inline - Subtraction

| | |
|---|---|
| Subtract | Off |
| Measurements | 1 |
| StdDev | Off |
| Save Original Images | On |

#### Inline - MIP

| | |
|---|---|
| MIP Sag | Off |
| MIP Cor | Off |
| MIP Tra | Off |
| MIP Time | Off |
| Radial MIP | Off |
| Save Original Images | On |
| MPR Sag | Off |
| MPR Cor | Off |
| MPR Tra | Off |

#### Inline - Composing

| | |
|---|---|
| Inline Composing | Off |

#### Sequence - Part 1

| | |
|---|---|
| Sequence Name | spcR |
| Dimension | 3D |
| Excitation | Non-sel. |
| RF Pulse Type | Normal |
| Gradient Mode | Normal |
| Flow Compensation | None |
| Reordering | Linear |
| Bandwidth | 723 Hz/Px |
| Echo Spacing | 3.68 ms |
| Turbo Factor | 282 |
| Echo Train Duration | 931 ms |

#### Sequence - Part 2

| | |
|---|---|
| Introduction | On |

#### Sequence - Assistant

| | |
|---|---|
| SAR Assistant | Off |
| Allowed Delay | 30 s |

| \\MedPhys\ABI\Schwaderlapp\EASEE_Studie_v2\T2w_space_dark-fluid_sag_1mm_iso |
|---|
| TA: 6:32 min Coil Selection: Manual Voxel Size: 1.0×1.0×1.0 mm³ Acc:: 2 Rel. SNR: 1.00 |

#### Properties

| | |
|---|---|
| Start measurement without further preparation | On |
| Wait for User to Start | On |
| Start measurements | Single Measurement |
| Prio Recon | Off |
| Auto Open Inline Display | Off |
| Auto Close Inline Display | Off |
| Load Images to MR View&GO | On |
| Auto Store Images | On |
| Disable auto transfer to PACS | Off |
| Load Images to Stamp Segments | On |
| Load Images to Graphic Segments | Off |
| Graphic segment | Default |
| Inline Movie | Off |

#### Routine

| | |
|---|---|
| Slab Group | 1 |
| Slabs | 1 |
| Position | R8.0 A11.6 H7.8 mm |
| Orientation | S > C-17.3 > T-5.2 |
| Phase Encoding Dir. | A >> P |
| Slices per Slab | 176 |
| Phase Oversampling | 0 % |
| Slice Oversampling | 0.0 % |
| FOV Read | 256 mm |
| FOV Phase | 100.0 % |
| Slice Thickness | 1.00 mm |
| TR | 5000.0 ms |
| TE | 386.00 ms |
| Averages | 1.0 |
| Concatenations | 1 |
| AutoAlign | Head > Brain |
| Coil Elements | HC1-7;NC1,2 |

#### Contrast - Common

| | |
|---|---|
| TR | 5000.0 ms |
| TE | 386.00 ms |
| MTC | Off |
| Magn. Preparation | Non-sel. T2-IR |
| TI 1 | 1800 ms |
| Flip Angle Mode | T2 Var |
| Fat-Water Contrast | Standard |
| Dark Blood | Off |
| Blood Suppression | Off |
| Wrap-up Magn. | None |
| Reconstruction | Magnitude |

#### Contrast - Dynamic

| | |
|---|---|
| Dynamic Mode | Standard |
| Measurements | 1 |
| Multiple Series | Each Measurement |

#### Contrast - Dynamic

| | |
|---|---|
| Reordering | Linear |

#### Resolution - Common

| | |
|---|---|
| FOV Read | 256 mm |
| FOV Phase | 100.0 % |
| Slice Thickness | 1.00 mm |
| Base Resolution | 256 |
| Phase Resolution | 100 % |
| Slice Resolution | 100 % |
| Interpolation | Off |

#### Resolution - Acceleration

| | |
|---|---|
| Acceleration Mode | GRAPPA |
| Total Factor | 2 |
| Reference Scans | Integrated |
| Acceleration Factor PE | 2 |
| Reference Lines PE | 24 |
| Acceleration Factor 3D | 1 |
| Phase Partial Fourier | Allowed |
| Slice Partial Fourier | 7/8 |
| Elliptical Scanning | Off |

#### Resolution - Filter

| | |
|---|---|
| Raw Filter | On |
| Elliptical Filter | Off |
| Distortion Correction | 2D |
| Normalize | Prescan |
| Image Filter | Off |

#### Geometry - Common

| | |
|---|---|
| Slab Group | 1 |
| Slabs | 1 |
| Position | R8.0 A11.6 H7.8 mm |
| Orientation | S > C-17.3 > T-5.2 |
| Phase Encoding Dir. | A >> P |
| Slices per Slab | 176 |
| Phase Oversampling | 0 % |
| Slice Oversampling | 0.0 % |
| FOV Read | 256 mm |
| FOV Phase | 100.0 % |
| Slice Thickness | 1.00 mm |
| TR | 5000.0 ms |
| Concatenations | 1 |

#### Geometry - AutoAlign

| | |
|---|---|
| Slab Group | 1 |
| Position | R8.0 A11.6 H7.8 mm |
| Orientation | S > C-17.3 > T-5.2 |
| Phase Encoding Dir. | A >> P |
| AutoAlign | Head > Brain |

#### Geometry - AutoAlign

| | |
|---|---|
| Initial Position | Isocenter |
| R | 0.0 mm |
| A | 0.0 mm |
| H | 0.0 mm |
| Initial Orientation | Sagittal |
| Initial Rotation | 0.00 deg |

#### Geometry - Navigator

#### Geometry - Saturation

| | |
|---|---|
| Special Saturation | None |

#### Geometry - Tim Planning Suite

| | |
|---|---|
| Set-n-Go Protocol | Off |
| Table Position | 0 mm |
| Table Position | H |
| Inline Composing | Off |

#### System - Miscellaneous

| | |
|---|---|
| Coil Selection | Manual |
| Radial Sorting | Off |
| MSMA | S - C - T |
| Sagittal | R >> L |
| Coronal | A >> P |
| Transversal | F >> H |
| Coil Combination | Adaptive Combine |
| Matrix Optimization | Off |

#### System - Adjustments

| | |
|---|---|
| Adjustment Strategy | Standard |
| B0 Shim | Standard |
| B1 Shim | TrueForm |
| Adjustment Tolerance | Auto |
| Adjust with Body Coil | Off |
| Confirm Frequency | Never |
| Assume Silicone | Off |

#### System - Adjust Volume

| | |
|---|---|
| Position | R8.0 A11.6 H7.8 mm |
| Orientation | S > C-17.3 > T-5.2 |
| Rotation | -19.57 deg |
| A >> P | 256 mm |
| F >> H | 256 mm |
| R >> L | 176 mm |
| Reset | Off |

#### System - pTx

| | |
|---|---|
| B1 Shim | TrueForm |
| Excitation | Non-sel. |

#### System - Tx/Rx

| | |
|---|---|
| Frequency 1H | 123.194104 MHz |
| ? Ref. Amplitude 1H | 0.000 V |
| Reset | Off |

#### System - Tx/Rx

| | |
|---|---|
| Correction Factor | 1.00 |
| Image Scaling | 1.000 |
| Gain | High |

#### Physio - Signal

| | |
|---|---|
| 1st Signal/Mode | None |
| Trigger Delay | 0 ms |
| TR | 5000.0 ms |
| Concatenations | 1 |

#### Physio - Cardiac

| | |
|---|---|
| Fat-Water Contrast | Standard |
| Magn. Preparation | Non-sel. T2-IR |
| TI 1 | 1800 ms |
| Dark Blood | Off |
| FOV Read | 256 mm |
| FOV Phase | 100.0 % |
| Phase Resolution | 100 % |

#### Physio - PACE

| | |
|---|---|
| Resp. Control | Off |
| Concatenations | 1 |

#### Inline - Subtraction

| | |
|---|---|
| Subtract | Off |
| Measurements | 1 |
| StdDev | Off |
| Save Original Images | On |

#### Inline - MIP

| | |
|---|---|
| MIP Sag | Off |
| MIP Cor | Off |
| MIP Tra | Off |
| MIP Time | Off |
| Radial MIP | Off |
| Save Original Images | On |
| MPR Sag | Off |
| MPR Cor | Off |
| MPR Tra | Off |

#### Inline - Composing

| | |
|---|---|
| Inline Composing | Off |

#### Sequence - Part 1

| | |
|---|---|
| Sequence Name | spcir |
| Dimension | 3D |
| Excitation | Non-sel. |
| RF Pulse Type | Normal |
| Gradient Mode | Normal |
| Flow Compensation | None |
| Reordering | Linear |
| Bandwidth | 751 Hz/Px |
| Echo Spacing | 3.68 ms |
| Turbo Factor | 278 |

#### Sequence - Part 1

| | |
|---|---|
| Echo Train Duration | 894 ms |

#### Sequence - Part 2

| | |
|---|---|
| Introduction | On |

#### Sequence - Assistant

| | |
|---|---|
| SAR Assistant | Off |
| Allowed Delay | 30 s |

| \\MedPhys\ABI\Schwaderlapp\EASEE_Studie_v2\fMRI_ME_TR1s_3mm_10min |
|---|
| TA: 10:23 min Coil Selection: Manual Voxel Size: 3.0×3.0×3.0 mm³ Acc:: 2 Rel. SNR: 1.00 |

#### Properties

| | |
|---|---|
| Start measurement without further preparation | On |
| Wait for User to Start | On |
| Start measurements | Single Measurement |
| Prio Recon | Off |
| Auto Open Inline Display | Off |
| Auto Close Inline Display | Off |
| Load Images to MR View&GO | On |
| Auto Store Images | On |
| Disable auto transfer to PACS | Off |
| Load Images to Stamp Segments | Off |
| Load Images to Graphic Segments | Off |
| Graphic segment | Default |
| Inline Movie | Off |

#### Routine

| | |
|---|---|
| Slice Group | 1 |
| Slices | 48 |
| Distance Factor | 0 % |
| Position | R5.4 A19.9 H8.2 mm |
| Orientation | T > C20.2 > S-1.1 |
| Phase Encoding Dir. | A >> P |
| Phase Oversampling | 0 % |
| FOV Read | 210 mm |
| FOV Phase | 100.0 % |
| Slice Thickness | 3.0 mm |
| TR | 1000.0 ms |
| TE 1 | 13.20 ms |
| TE 2 | 30.48 ms |
| TE 3 | 47.74 ms |
| Averages | 1 |
| Multi-band accel. factor | 4 |
| AutoAlign | Head > Brain |
| Coil Elements | HC1-7;NC1,2 |

#### Contrast - Common

| | |
|---|---|
| TR | 1000.0 ms |
| TE 1 | 13.20 ms |
| TE 2 | 30.48 ms |
| TE 3 | 47.74 ms |
| MTC | Off |
| Magn. Preparation | None |
| Flip Angle | 60 deg |
| Fat-Water Contrast | Fat Saturation |
| Contrasts | 3 |
| Reconstruction | Magn./Phase |

#### Contrast - Dynamic

| | |
|---|---|
| Dynamic Mode | Standard |
| Measurements | 600 |
| Delay in TR | 0.00 ms |

#### Resolution - Common

| | |
|---|---|
| FOV Read | 210 mm |
| FOV Phase | 100.0 % |
| Slice Thickness | 3.0 mm |
| Base Resolution | 70 |
| Phase Resolution | 100 % |
| Interpolation | Off |

#### Resolution - Acceleration

| | |
|---|---|
| Acceleration Mode | GRAPPA |
| Reference scan mode | GRE |
| Acceleration Factor PE | 2 |
| Reference Lines PE | 24 |
| Phase Partial Fourier | 7/8 |

#### Resolution - Filter

| | |
|---|---|
| Raw Filter | Off |
| Elliptical Filter | Off |
| Hamming | Off |
| Distortion Correction | Off |
| Static Field Correction | Off |
| Normalize | Off |

#### Geometry - Common

| | |
|---|---|
| Slice Group | 1 |
| Slices | 48 |
| Distance Factor | 0 % |
| Position | R5.4 A19.9 H8.2 mm |
| Orientation | T > C20.2 > S-1.1 |
| Phase Encoding Dir. | A >> P |
| Phase Oversampling | 0 % |
| FOV Read | 210 mm |
| FOV Phase | 100.0 % |
| Slice Thickness | 3.0 mm |
| TR | 1000.0 ms |
| Multi-Slice Mode | Interleaved |
| Series | Interleaved |
| Multi-band accel. factor | 4 |

#### Geometry - AutoAlign

| | |
|---|---|
| Slice Group | 1 |
| Position | R5.4 A19.9 H8.2 mm |
| Orientation | T > C20.2 > S-1.1 |
| Phase Encoding Dir. | A >> P |
| AutoAlign | Head > Brain |
| Initial Position | L0.0 A8.0 H3.2 |
| L | 0.0 mm |
| A | 8.0 mm |
| H | 3.2 mm |
| Initial Orientation | Transversal |
| Initial Rotation | 0.00 deg |

#### Geometry - Saturation

| | |
|---|---|
| Special Saturation | None |

#### Geometry - Tim Planning Suite

| | |
|---|---|
| Set-n-Go Protocol | Off |
| Table Position | 0 mm |
| Table Position | H |
| Inline Composing | Off |

#### System - Miscellaneous

| | |
|---|---|
| Coil Selection | Manual |
| Radial Sorting | Off |
| MSMA | S - C - T |
| Sagittal | R >> L |
| Coronal | A >> P |
| Transversal | F >> H |
| Coil Combination | Sum of Squares |
| Matrix Optimization | Off |

#### System - Adjustments

| | |
|---|---|
| Adjustment Strategy | Standard |
| B0 Shim | Brain |
| B1 Shim | TrueForm |
| Adjustment Tolerance | Auto |
| Adjust with Body Coil | Off |
| Confirm Frequency | Never |
| Assume Silicone | Off |

#### System - Adjust Volume

| | |
|---|---|
| Position | R5.4 A19.9 H8.2 mm |
| Orientation | T > C20.2 > S-1.1 |
| Rotation | -17.96 deg |
| A >> P | 210 mm |
| R >> L | 210 mm |
| F >> H | 144 mm |
| Reset | Off |

#### System - pTx

| | |
|---|---|
| B1 Shim | TrueForm |
| Excitation | Standard |

#### System - Tx/Rx

| | |
|---|---|
| Frequency 1H | 123.194104 MHz |
| ? Ref. Amplitude 1H | 0.000 V |
| Reset | Off |
| Correction Factor | 1.00 |
| Image Scaling | 1.000 |

#### Physio - Signal

| | |
|---|---|
| 1st Signal/Mode | None |
| TR | 1000.0 ms |
| Multi-band accel. factor | 4 |

#### BOLD

| | |
|---|---|
| GLM Statistics | Off |

#### BOLD

| | |
|---|---|
| Ignore Meas. at Start | 0 |
| Ignore After Transition | 0 |
| Model Transition States | On |
| Temp. Highpass Filter | On |
| Threshold | 4.00 |
| Paradigm Size | 20 |
| Meas[1] | Baseline |
| Meas[2] | Baseline |
| Meas[3] | Baseline |
| Meas[4] | Baseline |
| Meas[5] | Baseline |
| Meas[6] | Baseline |
| Meas[7] | Baseline |
| Meas[8] | Baseline |
| Meas[9] | Baseline |
| Meas[10] | Baseline |
| Meas[11] | Active |
| Meas[12] | Active |
| Meas[13] | Active |
| Meas[14] | Active |
| Meas[15] | Active |
| Meas[16] | Active |
| Meas[17] | Active |
| Meas[18] | Active |
| Meas[19] | Active |
| Meas[20] | Active |
| Motion Correction | Off |
| Spatial Filter | Off |
| Measurements | 600 |
| Delay in TR | 0.00 ms |

#### Sequence - Part 1

| | |
|---|---|
| Sequence Name | epfid |
| Dimension | 2D |
| Excitation | Standard |
| Gradient Mode | Normal |
| Flow Compensation | None |
| Bandwidth | 2380 Hz/Px |
| Echo Spacing | 0.52 ms |
| Free Echo Spacing | Off |
| EPI Factor | 70 |

#### Sequence - Part 2

| | |
|---|---|
| Introduction | On |
| RF Spoiling | Off |

#### Sequence - Special

| | |
|---|---|
| Excite pulse duration | 6000 us |
| Min. prep scans | 0 |
| Min. prep scans SB | 0 |
| iPAT ref. prep scans | 0 |
| Inter-TE delay | 0 us |
| Delay before PC scans | 0 us |
| Single-band images | On |

#### Sequence - Special

| | |
|---|---|
| MB LeakBlock kernel | On |
| MB dual kernel | Off |
| MB RF phase scramble | Off |
| Opt. MB RF pulse BW | Off |
| SENSE1 coil combine | Off |
| Invert RO/PE polarity | Off |
| Force Maxwell corr. | Off |
| PF omits higher k-space | Off |
| Disable freq. update | Off |
| Suppress 16-bit DICOM | Off |
| Force equal slice timing | Off |
| Online multi-band recon. | Online |
| FFT scale factor | 1.00 |
| Fat saturation FA | 110.00 deg |
| Fat sat. offset | 0.00 Hz |
| GRE iPAT ref. FA | 12.00 deg |
| Sinc exc. pulse BWTP | 5.20 |
| Physio recording | Legacy |
| Triggering scheme | Standard |

#### Sequence - Assistant

| | |
|---|---|
| SAR Assistant | Off |
