## Supplementary Methods 2 for "Post-implantation 3-T MRI in patients with an epicranial stimulation device: image quality and deep learning-based electrode reconstruction"

**Supplementary Methods 2. MRI-only electrode reconstruction**

The reconstruction pipeline used extracranial signal voids on post-implantation T1-weighted MPRAGE to estimate the position and orientation of the EASEE electrode contacts. Training examples were generated from MRI scans of five healthy participants by simulating 2,000 implant configurations per participant on skull surfaces segmented using SimNIBS. Simulations varied implant location, orientation, and distance from the skull and included the five-contact electrode geometry, connecting cable, and associated signal loss. Additional simulated scalp EEG contacts provided examples of unrelated signal voids. The known simulated electrode positions supplied the reference labels, avoiding the need for manually annotated patient scans. Data were separated by participant: four anatomies were used for training and the fifth for validation and model selection.

Reconstruction proceeded in two stages. First, two-dimensional views representing different depths beneath the head surface were generated from the MRI. A segmentation network (2D U-Net) identified the likely implant region in these views. This location was then used to extract an oriented three-dimensional MRI subvolume, in which a second network (3D U-Net) segmented the electrode contacts.

A template representing the known electrode geometry was subsequently aligned to the three-dimensional segmentation, first as a complete assembly and then with individual adjustments to each contact's position and orientation. The resulting contact masks were mapped back to the original MRI for comparison with the patient's cortical anatomy. The trained networks were applied to patient scans without further training or fine-tuning.

Further details are provided in the accompanying GitHub repository:
https://github.com/N-Schwaderlapp/EASEE_localization
